# Health system intervention packages on improving coverage of kangaroo mother care for preterm or LBW infants: a mixed-methods systematic review

**DOI:** 10.1101/2023.05.16.23289958

**Authors:** Nils Bergman, Megan Talej, Emily R. Smith, Suman PN Rao, Shuchita Gupta

## Abstract

**Introduction:** Global coverage of Kangaroo mother care (KMC) remains low and health system intervention strategies that may improve coverage are not known.

**Methods:** We conducted a systematic review of studies evaluating the effect of health system intervention strategies for KMC implementation compared to no or different interventions, on KMC coverage in preterm or LBW infants. KMC coverage achieved by various studies was summarized. All included studies were classified as those that achieved increased KMC coverage (defined as ≥25% increase from baseline, with final coverage ≥50%) or low KMC coverage (defined as <25% increase from baseline or final coverage <50%). Studies that achieved increased KMC coverage were further classified based on the mean duration of skin-to-skin contact (SSC; hours per day) achieved. Health system interventions in different categories were summarized by WHO health system building blocks to understand factors linked to increased KMC coverage.

**Findings:** We identified 16 studies evaluating 15 health system intervention packages for KMC implementation that applied interventions in one or more health system building blocks that reported KMC coverage. All three studies that applied interventions across 5-6 building blocks (100%), two of the four studies that applied interventions across 3-4 building blocks (50%), and three of the nine studies that applied interventions across 1-2 building blocks (33%), achieved increased KMC coverage. Studies that did not achieve increased coverage had interventions primarily targeting health workforce and service delivery and were weak on leadership and governance, financing, and health information systems. All three studies that achieved increased KMC coverage with mean SSC ≥8h/d (100%), three of the five studies that achieved increased KMC coverage with mean SSC <8h/d (60%), and three of the eight studies with low KMC coverage (38%) had high-intensity interventions in at least one health system building blocks. High-level leadership engagement, KMC supportive policies, staff licensing, and facility standards regulations, strengthened numbers and capacity of nursing staff, government funding and expanded health insurance, wards with conducive environment, and recording KMC-specific indicators in clinical registers were key factors among studies that achieved increased KMC coverage.

**Conclusion:** High-intensity interventions across multiple health system building blocks should be used for equitable scale-up of KMC.

## INTRODUCTION

Infants with low birth weight (LBW, birth weight below 2500 g) who are born preterm (gestational age at birth <37 weeks), are small for their gestational age (<10^th^ percentile for gestational age), or both, constitute approximately 15% of all neonates worldwide but account for 70% of all neonatal deaths (1,2). Kangaroo mother care (KMC) for preterm or LBW infants, defined as continuous and prolonged skin-to-skin contact of the infant with the chest of the mother (or another caregiver when not possible with the mother) and exclusive breastfeeding or breastmilk feeding, is a high-impact intervention that reduces neonatal mortality by 32% (3,4). Taken to scale, it has the potential to have a major impact on neonatal mortality and is a public health imperative.

There are few published data on population-based coverage of KMC, but it is known that the global KMC coverage remains low despite the long-standing WHO guidelines, country-level policies, and continued advocacy and efforts by global organizations (5). In 2019, only 32% of the 90 countries that reported progress on Every Newborn Action Plan had an updated policy or guideline on KMC (6). Even in countries with policies or guidelines on KMC, these are often not translated into programmatic implementation and many implementation barriers and potential facilitators for implementing KMC have been reported from different contexts. However, these are mostly based on multi-stakeholder consultations, expert opinions, or parents’ or health providers’ perspectives (7-9). While useful to understand these aspects, it is important to also look at the evidence of what strategies have worked in improving KMC coverage in real-life settings when the proposed solutions are operationalized.

However, there has been no systematic review of evidence on how to take KMC to scale. Health system interventions to improve implementation and achieve high population-based coverage of KMC in infants born preterm or low birth weight are not known. Therefore, we undertook the current review to understand which health system intervention strategies for KMC implementation increase its coverage in preterm or LBW infants.

## METHODS

The protocol for this systematic review was developed according to the Preferred Reporting Items for Systematic Review and Meta-Analysis Protocols (PRISMA-P) statement and registered with the PROSPERO International Prospective Register of Systematic Reviews (PROSPERO CRD42021271834) (10).

### Type of studies

We included interventional studies - randomized trials (individually randomized and cluster-randomized), non-randomized trials, before-and-after studies, interrupted time series, and repeated measure studies that evaluated the effect of health system interventions for KMC implementation meeting the quality criteria used by the Cochrane Effective Practice and Organisation of Care (EPOC) Group (11), applied in one or more of the WHO health systems building blocks, compared to no or different (non-health system) interventions, on KMC coverage in preterm or LBW infants. However, because of the paucity of studies, we also included uncontrolled before-and-after studies, which is a departure from the EPOC criteria.

All studies that reported an intervention package directed at improving KMC coverage in facilities or communities irrespective of baseline KMC coverage and effect were considered for inclusion. We excluded studies that only sought to improve the duration of KMC and/or encourage earlier initiation.

### Participants/population

Participants were preterm (born at <37 completed weeks of gestation) or LBW (birth weight <2500g) infants.

### Interventions

We included studies that applied interventions in one or more health system building blocks for improving KMC implementation. KMC was defined as continuous and prolonged skin-to-skin contact (SSC) of the infant with the chest of the mother (or another caregiver), feeding exclusively with breast milk, with or without early discharge from the hospital for preterm or LBW infants (12,13). We included all studies that reported ‘any KMC’ i.e., irrespective of SSC duration, support for exclusive breastfeeding or breastmilk feeding or early discharge, as long as it was clear that the intention was to provide continuous and prolonged SSC beyond the first hour after birth. Studies which focused only on SSC in the first hour after birth for all or term normal birth weight infants were excluded.

The interventions could have been applied at any scale or health system level, e.g., facility, home or community, or district or national levels, and could include one or more health system components to improve KMC practice. The health system actions were defined using the WHO health system building blocks framework, encompassing the domains of leadership and governance, health financing, health workforce, equipment and supplies, commodities, infrastructure, and service delivery, and health information systems (14).

### Comparator

Comparison groups were those that received no intervention to improve KMC or a different package of interventions, i.e., not targeting any health system components defined above.

### Outcomes

The primary outcome was coverage of “any KMC”, i.e., the proportion of eligible preterm or LBW infants who received ‘any KMC’ as defined above. The secondary outcome was the description of the components of the packages of health system intervention strategies that achieved increased (≥50%) coverage of ‘any KMC’.

### Search strategy

We systematically searched MEDLINE, Ovid, WHO Global Index Medicus, CINAHL, SCOPUS, Epistemonikos, and Web of Science, with no limit on language or date. The search was not limited by language, and all the potentially eligible studies were translated, if required, for inclusion in the review. The search was first done in August 2021, and updated in June 2022. Two authors (NB & MT) screened titles and abstracts using Covidence software and then undertook full-text review for assessing the eligibility of the studies. Any conflicts were resolved by two other authors (SG & SR). The search strategy is available on the link: https://www.crd.york.ac.uk/PROSPEROFILES/271834_STRATEGY_20210804.pdf

### Risk of bias in included studies

We assessed the risk of bias using RoB 2.0 for randomized trials and ROBINS-I for non-randomized studies (15,16)..

### Data extraction, analysis, and interpretation

Two authors extracted study details (MT, NB) and one author extracted numerical data for analysis (SG) which was independently cross verified by a second author (SR). The coverage of any KMC at baseline and end line was summarized for all studies, including the change in coverage achieved. The eligible studies were too heterogeneous to be pooled and hence a meta-analysis was not conducted. The quantitative synthesis of KMC coverage was followed by qualitative analysis using a sequential/consecutive approach for mixed-methods systematic reviews (17-19). All eligible studies were classified based on the final KMC coverage achieved and change in KMC coverage from baseline, as follows:

a. Studies that achieved increased KMC coverage (defined as a ≥25% increase from baseline, with final coverage ≥50%); or
b. Studies that achieved low KMC coverage (defined as <25% increase from baseline or final coverage <50%)

Studies that achieved increased KMC coverage were further classified based on the mean duration of SSC (hours per day) achieved, as shown in table 1.

**Table 1.**
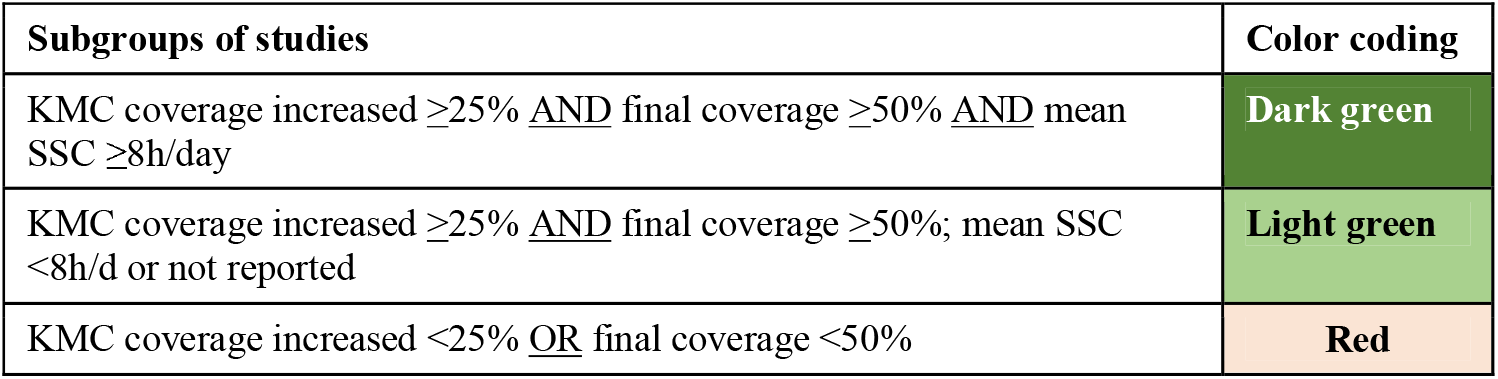
Subgrouping of studies using quantitative results on KMC coverage

| Subgroups of studies | Color coding |
| --- | --- |
| KMC coverage increased $\geq 25\%$ <u>AND</u> final coverage $\geq 50\%$ <u>AND</u> mean SSC $\geq 8\text{h/day}$ | Dark green |
| KMC coverage increased $\geq 25\%$ <u>AND</u> final coverage $\geq 50\%$; mean SSC $< 8\text{h/d}$ or not reported | Light green |
| KMC coverage increased $< 25\%$ <u>OR</u> final coverage $< 50\%$ | Red |

Thereafter, the interventions applied by the different studies were classified using the WHO health system building blocks framework as in Table 2 (14).

**Table 2.**
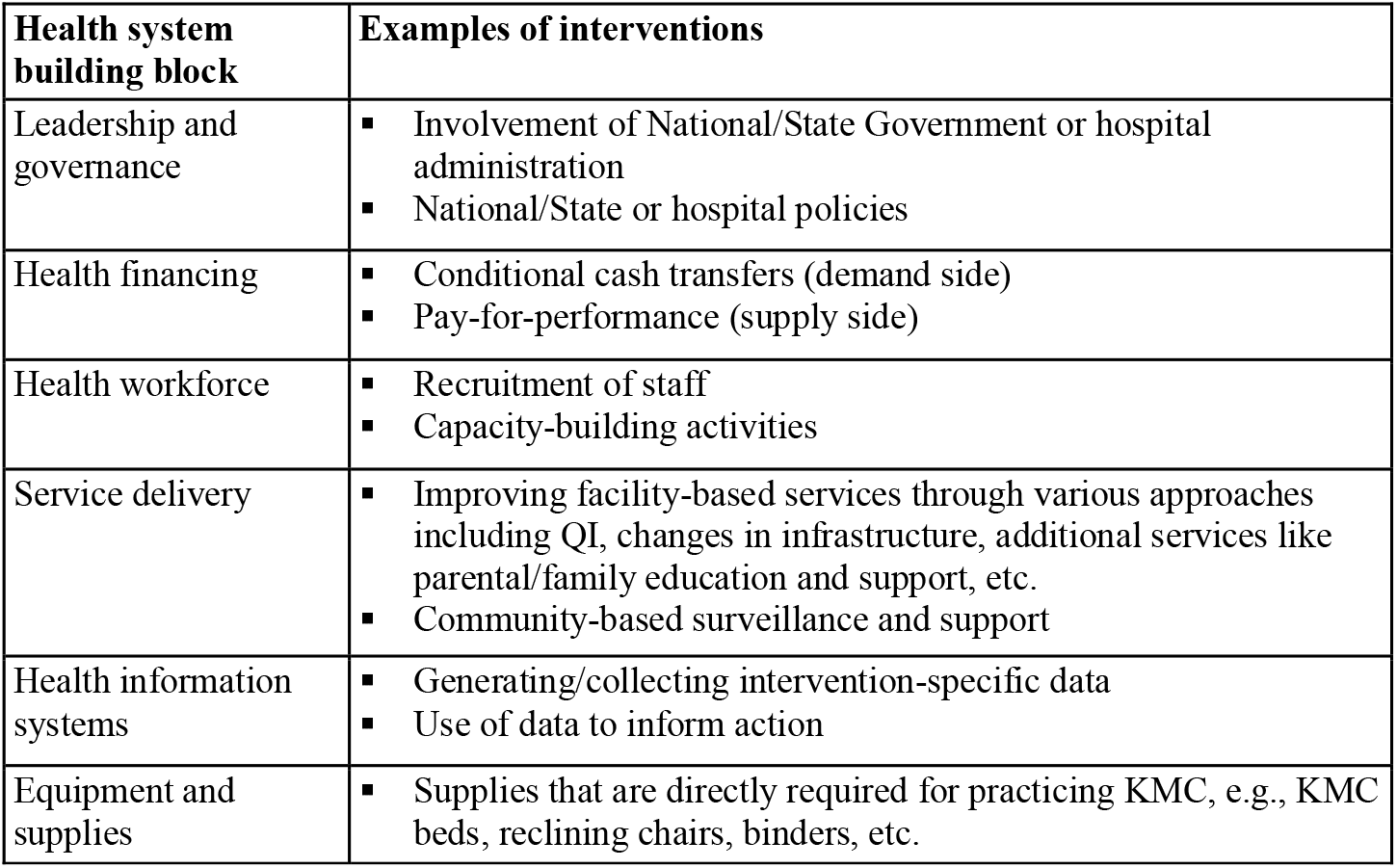
Classification used for health system interventions for KMC implementation

We considered the number of health system building blocks in which the interventions were applied as the “breadth” of intervention. Based on the six health system building blocks described above, we considered the “breadth” of intervention as high if a study intervention package comprised interventions in 5 or 6 building blocks, moderate if it applied interventions in 3 or 4 building blocks, and low if the package included interventions only in 1 or 2 health system building blocks.

We also evaluated the intensity of health systems interventions for KMC implementation using Cochrane standard approach for assessing the intensity of complex interventions, using the TIDieR criteria (20), as: 1) How much-Number of times and over what period intervention was delivered, including the number of sessions, their schedule, duration, intensity, or dose; and 2) How well-If intervention adherence or fidelity was assessed, the extent to which the intervention was delivered as planned; whether or not any strategies were used to maintain or improve fidelity. Two independent assessors (SG, SR) categorized each intervention in each health system building block as high or low intensity. The intervention was marked as high if both reviewers considered it as high, else it was marked as a low-intensity intervention.

The ‘breadth’ and ‘intensity’ of interventions were compared and contrasted between the different categories of studies based on the KMC coverage categories to understand the factors that may lead to high coverage of any KMC.

## RESULTS

We identified 16 studies that evaluated 15 health system intervention strategies for KMC implementation applied in one or more building blocks and reported KMC coverage (Figure 1). Two studies implemented the same intervention package in two countries, Uganda, and Nepal, resulting in two publications.

**Figure 1.**
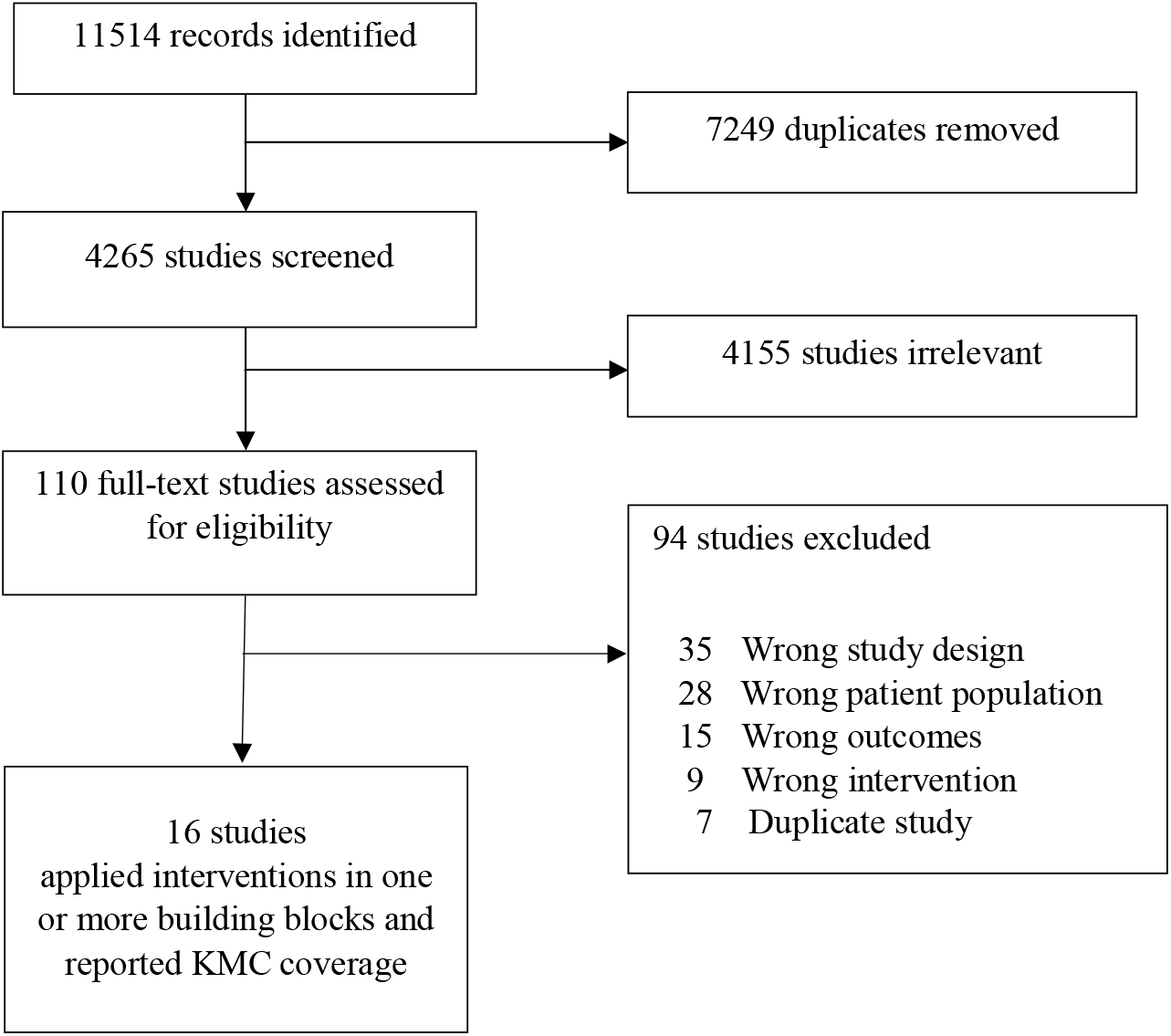
PRISMA flow chart.

### Design and setting

Two studies were community-based cluster randomized trials (21,22) two were reports of a national-level programmatic scale-up (23,24), one was a mixed-methods study across facility-community continuum at district/woreda level (25), three were quality-improvement studies – one facility and one community-based (26-28), and eight were uncontrolled before-after health facility-based studies (28-35). Thirteen studies were conducted in lower-middle-income or low-income country settings -Bangladesh, Ethiopia, Ghana, India, Nepal, Philippines, Uganda) and three in high-income settings (USA) (Table 3).

**Table 3.** Characteristics of included studies (n=16)

| Study Country | Study setting and methods | Participants | Intervention details | Outcome reported | Notes |
| --- | --- | --- | --- | --- | --- |
| Arora 2021, India (26) | Hospital-based pre- and post-intervention study conducted over 8 months (4 months baseline, 3 months intervention, and one-month sustenance phase). | PT or BW <2.5 kg clinically stable twins; a total of 169 twin pairs included in the study. | Intervention phase comprised three Plan-Do-Study-Act (PDSA) cycles.<br>The first cycle focused on counselling sessions and encouragement of family participatory care, KMC care was encouraged by allowing an additional caretaker at all times of the day<br>the second cycle comprised distribution of low-cost KMC bags and increasing bed strength in the KMC ward by redesigning two additional maternity wards and making them function as KMC wards, entry of fathers inside the KMC wards which was restricted as per hospital policy was relaxed and they were allowed to participate in KMC even during night<br>in the third cycle, dedicated KMC nurses were appointed- they were relieved from other duties. They counselled for family participated care with emphasis on early initiation, continuous KMC, positioning of twins, expression of breastmilk and feeding of LBW neonates, assisted mothers and families in practicing KMC, and maintained records mentioning day of initiation, KMC hours<br>KMC definition: SSC several hours per day; EBF support provided | % of mothers who initiated KMC during facility stay<br>% mothers who gave prolonged KMC for $\geq 8$ h | Analysis for this paper included n=91 twin pairs i.e., n=64 included in the baseline and n=27 included in the sustenance phase. |
| Calibo 2021 Philippines (24) | Mixed-methods evaluation of nationwide scale-up of KMC in 2014-2019 comprising secondary data review and in-depth interviews with policymakers, programme managers and programme coordinators of hospitals | **2015:** 179 postpartum maternal interviews from a sample of 17 national and subnational hospitals<br>**2019:** 478 postpartum maternal interviews from a sample of | The interventions for strengthening KMC (2014-2019) built on policy and programmatic actions to strengthen essential intrapartum and newborn care (EINC) in the previous four years (2009-2013), summarized below:<br>**2009–2013: EINC strengthening**<br>Administrative order adopting new policies and protocol for essential newborn care.<br>EINC practice protocol including KMC for the care of small babies and newborn care clinical practice pocket guide (job aid)<br>EINC incorporated into preservice curricula and national licensure examinations. | % infants <200 g/<36 weeks who received KMC | Only the KMC-specific interventions between 2014-2019 were considered in the review |
|  |  | 45 national and subnational hospitals | Reimbursement for newborn care package<br>EINC included in licensing standards for health facilities.<br>EINC included in medium/long-term development plans and budget allocated for national and local programmes<br>Communication campaigns on EINC<br>**2014-2019: KMC scale up**<br>Care for the small baby programme established with bilateral and multilateral partners and the KMC Foundation (2014)<br>Social health insurance and benefits for women about to give birth (2014)<br>the Z benefits for premature and small newborns (2017)<br>Enhancement of newborn care package (2018)<br>KMC definition: SSC and exclusive BF |  |  |
| Hendricks 2014<br>USA (31) | Hospital-based study; prospective pre- and post-study with 6 months pre-, 1-month intervention and 18 months-post-intervention period | Preterm infants $\leq 34$ weeks' gestation born in the hospital, n=112 (34 in pre 78 in post-intervention period) | The one-month KMC education and training program intervention comprised:<br>Three 7.5-hour sessions of didactic education on KMC<br>Five manikin KMC simulation practicums with infant mannequins receiving (1) room air, (2) nasal cannula, (3) nasal continuous positive airway pressure (NCPAP), (4) synchronized inspiratory positive airway pressure (SIPAP), and (5) intubation on conventional ventilators, followed by debriefing.<br>Nurses undergoing competency/skills training (n=30) were under direct observation in the NICU by senior staff every day for 6 months post-training<br>KMC definition: SSC, EBF support not specified | % of inborn infants $\leq 34$ weeks who were provided KMC during the pre- and post-training study period | None |
| Hodgins 2020<br>Nepal (22) | Community-based cluster-randomized trial to determine whether family-administered screening with targeted messages improves care practices | Women recruited late in pregnancy; 17 clusters in intervention and comparison arms each; Analysis based on low-birth-weight infants $< 2500$ g; n=1152 | Women were given a 6.9 cm card to assess whether the baby's foot is small; if so, to call a pre-provided number to listen to 3 cognitive and 2 affective pre-recorded messages. Follow-up visits were made over the 2 weeks following the birth within 24 h of birth (for health facility births, the first postnatal home visit could be on day 3 or later) and then on days 3, 7 10, and 14. The control group also received a basic set of interventions/support, but not the foot measurement card.<br>KMC definition: SSC, EBF support not specified | % LBW (BW$<2500$ g) infants who received SSC in first 14 days after birth | Infants were determined to be low birth weight based on birth weight taken by the field staff, and foot length $\leq 6.9$ cm, as determined by the family and by the field staff. |
| Joshi 2022<br>India (28) | Hospital-based study using quality improvement methods with multiple Plan-Do-Study-Act (PDSA) cycles over 12 weeks | All admitted stable LBW babies weighing less than 2500 g, numbers not reported. | KMC chairs and garments (front open gowns and binders), designated KMC block established, service delivery adaptations to minimise interruptions.<br>KMC definition: SSC, EBF support not specified | % of LBW babies who received KMC for at least 6 hours per day | Results reported only in percentage, denominator assumed to be 100 for baseline as well as sustenance phase.<br>Baseline data was recorded over seven consecutive days; final coverage data noted as a monthly average at the end of 26 weeks of the sustenance phase. |
| Kabir 2022<br>Bangladesh (23) | Evaluation of nationwide scale-up of KMC in government-run healthcare facilities in rural and urban areas of Bangladesh from Jan 2016 to Mar 2020 using data from the national database. | Clinically stable babies with birth weight under 2000 g, n=5018 | Government introduced KMC nationwide as a part comprehensive strategy to reduce neonatal mortality with defined coverage targets.<br>A dedicated space (corner) for KMC was established in each designated health facility i.e., subdistrict health complexes and higher-level facilities (district hospitals and medical college hospitals) in both urban and rural areas.<br>Government developed a national trainer pool for health-care professionals. Training programmes included a 3-day training focusing on practical demonstration, learning by doing, facility visits for direct observation of KMC services and recording and reporting of data.<br>A social and behavioural change communication campaign to create awareness in the community via printed materials (such as leaflets, posters, flipcharts and booklets), television, radio and social media platforms<br>Online KMC data set launched in 2016. Data entered by trained personnel in facilities and periodically monitored by the Government for completeness and accuracy. Staff from the national newborn health programme evaluated KMC data every month and acted on underreporting facilities. KMC services were monitored quarterly by government personnel whereby professionals and<br>programme personnel visited facilities and conducted onsite monitoring.<br>KMC was introduced formally into the curricula of medical and nursing professionals. In parallel, various professional organizations of paediatricians, obstetricians, neonatologists, and nursing colleagues endorsed the universal practice of KMC.<br>KMC definition: SSC, EBF support not specified | % of LBW babies <2000 g who received KMC | Over the 4-year period 64 426 babies weighing under 2000 g were born in the eligible facilities. In this review, we compared data from first quarter of 2016 (Jan - Mar 2016) as baseline and first quarter of 2020 (Jan-Mar 2020) as endline. |
| Kapoor 2021<br>India (35) | Hospital-based, pre-post intervention study over 9 months with 3 months each of pre- intervention- and post intervention phase | Hemodynamically stable preterm neonates admitted to the NICU with a birth weight of <2000 g, n=180 | A KMC education protocol comprising one-to-one counseling of the mother and the residential elderly woman on the benefits and procedure of KMC in immediate postnatal period in labour room and at the time of initiation of KMC in the NICU. A 10 min video film regarding benefits and procedure of KMC was shown in the NICU; pictorial brochures regarding procedure and benefits of KMC were shown and given to the mothers.<br>Focused group discussion conducted once a week with mothers and the residential elderly woman to allay their fears, address their concerns and educate and motivate them. Other members of the family especially the residential elderly woman were also motivated and encouraged to give KMC. Faculty of the area checked and supervised and addressed the glitches/issues.<br>The daily treatment order included a prescription for KMC by the resident doctor. Staff nurse on duty recorded hours of KMC in the patient record file and a KMC register was also maintained to record hours of KMC done per shift. At discharge, mothers were given a small booklet and pen and they were asked to maintain a record of the KMC provided every day. During every follow-up visit one-to-one counseling of the mother and other family members was done to address their concerns and motivate them for KMC.<br>KMC definition: SSC for several hours in a day with EBF | % of eligible preterm infants receiving KMC during hospital stay | None |
| Minot 2021<br>USA (30) | Hospital-based study using quality improvement | Infants born <28 weeks' gestation | Previously established unit policies and guidelines were updated based on current evidence to facilitate early SSC | % of extremely preterm or LBW | None |
|  | methods with multiple Plan-Do-Study-Act (PDSA) cycles over two years (Jun 2017 to Dec 2019) with one year baseline (June 2017-2018) and one-and-a-half-year intervention (Jul 2018 to Dec 2019). | and <1000 g, n=52 | and widely disseminated and posted within the hospital. SSC Readiness Checklist was developed and nurses and parents were asked to complete it prior to the first SSC. The checklist included guidance about duration of SSC and specific ways to prepare the parent, the environment, and the infant.<br>Education on evidence for the benefits of KMC was provided to health providers at staff meetings and specific small baby education days and encouraged to be discussed by the team daily in rounds.<br>Parents were offered frequent education on the importance of SSC through information cards, family-centered daily rounds and practice parent-led transfer of baby with a mannequin with endotracheal tube and intravenous lines before their first-time holding.<br>KMC definition: SSC ≥ 1 h/d | infants who received SSC within 72 h after birth |  |
| Mondkar 2021<br>India (29) | Hospital-based study pre- and post- intervention study with six months pre-(Jan-Jun 2017), 8 months intervention (Jul 2017-Feb 2018) and 6 months post-interventions (Mar-Aug 2018). Mothers with healthy term neonates in the postnatal care (PNC) wards and mothers with VLBW neonates admitted to the NICU were enrolled | Very low birth weight (VLBW) infants <1500 g at birth, n=227 | Multiple plan-do-study-act (PDSA) cycles were conducted simultaneously during the intervention period to improve clinical practices including KMC. Interventions for improving uptake of KMC included in-service training of healthcare providers by conducting an 8-h workshop with practical hands-on demonstration and strengthening counselling skills, doubling the number of KMC chairs to address shortage of chairs, positive reinforcement for best performing mothers, involving grandmothers in giving KMC and making KMC a monthly review indicator to track improvement.<br>KMC definition: SSC and EBF support provided | % of VLBW neonates receiving extended KMC | Extended KMC not defined |
| Mony 2021<br>India, Ethiopia (25) | Mixed-methods implementation research study in geographically defined rural and semi-urban study areas (one district in each site in India, 3–5 woredas (districts) in each site in Ethiopia). | Infants of birth weight <2000 g who survived the first 3 days, were available in the study area and whose mother resided in the study | The intervention models included three components:<br>pre-KMC facility activities aimed at maximizing access of LBW babies to KMC-implementing facilities; these activities included accurate birth weight recording and referral of LBW infants born at home or in facilities that did not provide KMC.<br>KMC-implementing facility activities aimed at initiating and maintaining KMC for all LBW babies weighing | % SSC ≥ 8 h per day before discharge | Study included three sites in India and four sites in Ethiopia, with some context-specific variations in the interventions across sites.<br>The study provided data |
| | Formative research 6 months (Mar-Sept 2016), three iterative cycles of intervention model development and refinement to continuously improve KMC coverage of 5-6 months each, and evaluation phase of 9—12 months (Jan 2018-Apr 2019) | area, n=3686 | <2000 g at birth who were born in or referred to the facility; these activities included changes in infrastructure and training, motivation, and support of facility staff.<br>3. Post KMC implementing facility activities aimed to support the continuation of KMC at home after discharge<br>KMC definition: SSC min 8 h/d, with EBF feeding support.<br>KMC definition: SSC for several hours in a day with EBF | | on additional outcomes like any SSC and SSC plus EBF $\geq 8$ h per day but we used the selected outcome to align with the rest of the studies. None of the hospitals in the study areas provided KMC at baseline, so baseline evaluation not done- denominator assumed to be the same as endline and KMC coverage assumed to be 5% as a conservative estimate. |
| Nation 2021 USA (34) | Hospital-based pre-post intervention study | Infants born before 29 weeks' gestation regardless of respiratory support; n=81. Infant with cardiovascular instability requiring inotropic administration and umbilical arterial catheters were excluded. | Newborn unit's existing SSC protocol was updated and modified to provide clarity on inclusion and exclusion criteria for SSC sessions, detailed description of transferring the newborn to and from the SSC position, and documentation. The electronic medical record was modified to include a place for bedside nursing to document the length of each SSC session and if any adverse events occurred.<br>The main intervention focused on an educational series provided directly to the nursing staff assigned to this population in the NICU and was tailored to overcome identified barriers and comprised both didactic sessions and simulation allowing for hands-on practice with transfer techniques for infants requiring intubation and CPAP respiratory equipment as well as IV and central lines. The educational sessions were offered over a 1-month period on 6 individual days and divided into 8 total sessions.<br>KMC definition: SSC, EBF support provided | % infants who received SSC within the first 7 days after birth | Initially, infants <26 weeks were excluded for the first 72 hours of life. During the project period this unit protocol changed to include any infant <26 weeks for first 14 days of age and infants 26 to 27 completed weeks for the first 7 days of age |
| Panda 2021 India (27) | Community-based study using QI methods with two | Singleton neonates with a discharge | Interventions during the first PDSA cycle included:<br>Demonstrate KMC to all eligible mothers and family | % of newborns receiving home- | The study implemented two PDSA cycles but it is |
|  | PDSA cycles over two months; n=29 eligible mother–neonate dyad One month (April 2020) baseline and two PDSA cycles (May-June 2020 and June to July 2020) | weight of < 1800 g singleton neonates born between April and July 2020 | members before discharge.<br>▪ Demonstrate KMC through videos.<br>▪ Provide one-to-one counseling over telephonic conversations.<br>▪ Create a WhatsApp group for all eligible discharged mothers.<br>▪ Interact through video calls with a female nurse educator.<br>Interventions during the second PDSA cycle included:<br>▪ Use KMC bag.<br>▪ Encourage family members to provide KMC.<br>▪ Announce KMC champion of the day.<br>KMC definition: Post-discharge SSC, Breastmilk express | based KMC | not clear whether interventions from the first PDSA cycle were continued during the second PDSA cycle or not. Therefore, we used data only from the first PDSA cycle for which complete information was available. |
| Prasantha 2019 India (36) | Hospital-based pre-post intervention study over five months with one month each of pre- and post- and three months of intervention period | Not specified/likely to be preterm or LBW infants <2000 g | ▪ KMC wards and lactation clinics established- mothers allowed inside SNCU for feeding and KMC.<br>▪ Training of health providers with on-going supervision and on-job support<br>▪ KMC audit<br>KMC definition: SSC and EBF support provided | % of eligible newborns initiated on KMC | The hospital follows Government guidelines wherein KMC is provided to stable preterm or LBW newborns <2000g |
| Spira 2017 Uganda (33) | Hospital-based pre-post intervention study over six months (Mar- Oct 2014). Women >18 years enrolled in two hospitals, n= 4816 | LBW newborns <2000g, n=56 (38 from one hospital and 18 from other) | Joint promotion by three healthcare professional associations in two hospitals' comprising six activities: dissemination workshops, development of reminders, delivery simulation sessions, team building, case reviews, and academic visits in the wards.<br>KMC definition: SSC and EBF support | % of preterm and/or low birthweight (BW<2000 g) neonates who received KMC | None |
| Spira 2018 Nepal (32) | Hospital-based pre-post intervention study over six months (Jun to Dec 2016) Women >18 years enrolled in two hospitals, n=9231 | Preterm or LBW newborns <2000g, n=254 in one hospital | Same as above<br>KMC definition: SSC and EBF support | % of preterm and/or low birthweight (BW<2000 g) neonates who received KMC | Data only from one hospital has been used because the denominator for post-intervention period was not available from the other hospital. |
| Vesel 2013 Ghana (21) | Cluster-randomized trial 49 intervention zones and 49 control zones, In seven districts in the Brong Ahafo Region, | Facility-born LBW babies with known facility birth weight <2.5 kg, n=745 | Existing community-based volunteers were trained to conduct home visits during which they weighed babies and counselled mothers of LBW babies on SSC in the intervention zones.<br>KMC definition: SSC | % of LBW newborns who received any SSC and SSC for more than 2 h. | Of 15,615 live births, 68.5% had recorded birthweights; 10.1% were LBW. |
|  | Ghana, including all live births between Nov 2008 and Dec 2009 |  |  |  |  |

**Table 4.** Any KMC coverage and change in KMC coverage achieved by included studies by KMC coverage categories

| KMC coverage categories | Study (author, year) | Any KMC coverage<br>(Proportion of eligible preterm or LBW infants who received any KMC) |  | Change in any KMC coverage from baseline | Mean skin-to-skin contact per day achieved |
| --- | --- | --- | --- | --- | --- |
|  |  | Intervention group | Comparison group |  |  |
| Coverage increase $\geq 25\%$ AND final coverage $\geq 50\%$ AND SSC $\geq 8$h/day | Mony 2021 (25) | 2344/3686 (64%)* | 0** | 64% | $\geq 8$ hours per day prior to discharge |
| | Arora 2021 (26) | 22/27 (82%) | 34/64 (53%) | 29% | $\geq 8$ hours per day prior to discharge |
| | Kapoor 2021 (35) | 92/92 (100%) | 66/88 (75%) | 25% | $\geq 8$ hours per day prior to |

|  |  |  |  |  | discharge |
| --- | --- | --- | --- | --- | --- |
| Coverage increase $\geq 25\%$ AND final coverage $\geq 50\%$ | Joshi 2022<sup>#</sup> (28) | 76/100 (76%) | 30/100 (30%) | 46% | 6 hours per day during hospital stay |
|  | Minot 2021 (30) | 31/37 (84%) | 1/15 (7%) | 77% | Not reported |
|  | Hendricks-Munoz 2014 (31) | 67/78 (86%) | 9/34 (26%) | 60% | About 4 hours per day during hospital stay |
|  | Calibo 2021 (24) | 253/478 (53%) | 19/179 (11%) | 42% | Not reported |
|  | Mondkar 2021 (29) | 81/114 (71%) | 49/113 (43%) | 28% | Not defined, but “extended KMC”, likely to be at least 6 hours per day during hospital stay |
| Coverage increase $< 25\%$ OR final coverage $< 50\%$ | Panda 2021 (27) | 5/10 (50%) | 3/8 (38%) | 12% | Not reported |
|  | Prashantha 2019<sup>#</sup> (36) | 42/100 (42%) | 2/100 (2%) | 40% | Not reported |
|  | Vesel 2013 (21) | 190/375 (51%) | 105/370 (28%) | 23% | Not reported |
|  | Hodgins 2020 (22) | 186/503 (37%) | 96/649 (15%) | 22% | Not reported |
|  | Nation 2021 (34) | 15/24 (63%) | 13/25 (52%) | 11% | Not reported |
|  | Kabir 2022 (23) | 917/4226 (22%) | 37/792 (5%) | 17% | Not reported |
|  | Spira 2018 (Nepal) (32) | 23/124 (19%) | 29/127 (23%) | 4% | Not reported |
|  | Spira 2017 (Uganda)<sup>##</sup> (33) | 12/20 (60%) | 29/36 (81%) | -21% | Not reported |

\* Combined data from seven different sites where coverage varied from 55% to 85%; \*\* None of the hospitals in the study areas provided KMC at baseline
<sup>#</sup> Study reported only in percentage, denominator assumed to be 100 for both groups
<sup>##</sup> Combined data from two hospitals

### Population

The included studies either enrolled the mothers before delivery and reported the outcomes among those that delivered preterm or LBW infants or enrolled preterm or LBW infants after birth. While all studies enrolled preterm or LBW infants, the eligibility criteria varied in terms of birth weight and gestational age across different studies (table 3).

### Interventions applied by the included studies

All studies applied interventions to improve KMC practice for preterm or LBW infants, at variable scale, depending on their setting, contextual factors and baseline KMC coverage/practice. While all studies did not report support for EBF/EBM feeding as part of the intervention package, it was provided as part of the unit protocol in many studies which also reported breastfeeding outcomes.

### Outcomes

All studies reported the proportion of eligible preterm or LBW infants who received skin-to-skin contact after birth in the pre-intervention phase or comparison group and post-intervention phase or intervention arm. However, the duration of skin-to-skin contact and the time of measurement varied (table 3).

There was high heterogeneity in the studies reporting KMC coverage in terms of population, implementation level and scale, KMC definition and reported outcomes, though all studies specified skin-to-skin contact more than one hour and the proportion of infants who received KMC. The risk of bias assessment for included studies and the list of excluded studies with reasons are provided in the Supplementary Appendix.

### Quantitative assessment of KMC coverage

Eight of the 16 studies achieved increased KMC coverage with final KMC coverage ≥50% and an increase of ≥25% from baseline (Table 3). Of these, three studies reported SSC duration ≥8 hours per day. One was large mixed methods implementation research study conducted in 8.7 million population in India and Ethiopia, that achieved a final KMC coverage (SSC for ≥8 hours per day) of 55-85% at different sites, with an increase in the population-based coverage of KMC by more than 75% across seven sites in the two countries. The mean duration of skin-to-skin contact was 9.6 to 12.0 h/d in India sites and 11.6 to 14.9 h/d in Ethiopia sites (table 3)

We reviewed the interventions applied by all included studies and classified them in various health system building blocks by study, based on pre-specified matrix provided in table 2 (table 5). The high intensity interventions are marked in bold font.

**Table 5.** Interventions applied by the included studies classified by health system building blocks*

| Study | Leadership and Governance | Health workforce | Service delivery | Supplies | Health information systems | Health financing |
| --- | --- | --- | --- | --- | --- | --- |
| Mony 2021 (25) | State/Provincial leadership strongly engaged<br>KMC supportive policies established (Mothers given rights and means to stay with their babies- beds, food, bathing, toilet, etc.) | Additional nurses appointed; personnel specially assigned to support KMC activities. HWs including CHWs trained, motivated, supported to perform the required tasks. | Activities in lower-level facilities and community to identify and refer LBW babies to KMC implementing facilities<br>KMC wards with conducive environment, counseling<br>Post-discharge phone calls and home visits to support continued KMC at home | KMC beds, reclining chairs | KMC-specific indicators incorporated into routine data systems. Registries and clinical logs developed to track small babies in health facilities | KMC costs budgeted by the Govt (wards, human resources including incentives to CHWs, running expenditures) |
| Arora 2021 (26) | Fathers and additional caregivers allowed to enter KMC wards and provide KMC 24x7 | Dedicated nurses appointed to KMC wards, training of staff in supporting KMC practice and counselling | Increased number of beds in KMC ward, curtains installed in KMC ward, counselling sessions and encouragement of family participatory care | Low-cost KMC garments | Records of day of initiation, KMC hours on daily basis |  |
| Kapoor 2021 (35) |  |  | KMC counseling to mothers and accompanying elderly woman, focused group discussions, other family members encouraged to give KMC. KMC in daily treatment order. |  | KMC record maintained in patient file and service register |  |
| Joshi 2022 (28) |  |  | Dedicated KMC block established, service delivery adaptations to minimize interruptions | KMC chairs, garments, binders |  |  |
| Minot 2021 (30) | Unit policies and guidelines on KMC updated on all aspects | Staff and provider education on evidence for SSC. | Parent education and practical skill training. Readiness checklist |  |  |  |
| Hendricks 2014 (31) |  | KMC education and training program with simulation-based training for nurses using high-fidelity mannequins, role play and didactic education |  |  |  |  |
| Calibo 2021 (24) | Regulations to support evidence-based policies around birth, updated training curricula, staff licensing standards, facility regulatory and standards. KMC combined with essential intrapartum and newborn care and resuscitation in one initiative | Engagement of professional associations<br>KMC 'champions', maternity - neonatal staff collaboration, for practice change, on-job clinical coaching | Environmental changes including reorganisation of space, patient flow, staff allocation, equipment and supplies, and education activities.<br>KMC promoted through multiple communication channels | KMC beds and binders | Self-monitoring by hospital teams and periodically by external teams at a national scale. | DoH committed financing for implementing KMC as part of essential intrapartum and newborn care by incorporating KMC in annual implementation plans. Expanded health insurance package for small babies. |
| Mondkar 2021 (29) |  | Training of health care providers with practical hands-on demonstration on KMC technique and strengthening counselling skills | Positive reinforcement for best performing mothers, involving grandmothers in giving KMC | KMC chairs increased | KMC made a monthly review indicator |  |
| Panda 2021** (27) |  |  | **Family counselling and support through demonstration, videos, telephonic counseling and video calls.** |  |  |  |
| Prasantha 2019 (36) | Mothers allowed inside SNCU for feeding and KMC | Training of health providers with on-going supervision and on-job support | KMC wards and lactation clinics established |  | KMC audit |  |
| Vesel 2013 (21) |  | Training of CHWs to promote SSC for LBW infants during home visits. | Mother shown photograph of SSC performed by a woman from their area, informed that healthcare providers recommend SSC for LBW babies. Family members encouraged to help mothers with SSC. |  |  |  |
| Hodgins 2020 (22) |  |  | Family-administered screening to identify LBW infants; toll-free numbers with pre-recorded phone messages including advice on skin-to-skin contact. |  |  |  |
| Nation 2021 (34) |  | **Simulation training with hands-on practice of nurses tailored to overcome identified barriers** | KMC protocol updated and modified to provide clarity on eligibility, details of SSC practice and documentation |  |  |  |
| Kabir 2022 (23) | **National strategy with defined coverage targets, facility/DHIS2 data periodically monitored by Government for completeness and accuracy, KMC introduced formally into the curricula of medical and nursing professionals** | National trainer pool for health-care professionals, quarterly on-site monitoring of KMC services, professional organizations endorsed universal practice of KMC | Social and behavioural change communication campaign to create awareness in the community |  | **Online KMC data set established – facility record-keeping by trained personnel with regular review and action on underreporting facilities.** |  |
| Spira 2017 (33) |  | KMC promotion through dissemination workshops, development of reminders, delivery simulation sessions, team building, case reviews, and academic visits in the wards. |  |  |  |  |
| Spira 2018 (32) |  | Same as above for Spira 2017 |  |  |  |  |

\*The text in bold indicates the interventions whose intensity was considered 'high'; \*\* Interventions only during the first PDSA cycle are included
Abbreviations; SNCU- Special newborn care unit, CHWs- community health workers, SSC- Skin-to-skin contact, EBF-Exclusive breastfeeding, LBW-Low birth weight

### Qualitative analysis of health system interventions by KMC coverage categories

A higher proportion of studies that applied interventions across more health system building blocks achieved increased coverage:

- All three studies (100%) that applied interventions across 5-6 building blocks achieved increased coverage: Mony 2021, Arora 2021 and Calibo 2021
- Two of the four studies (50%) that applied interventions across three to four building blocks achieved increased coverage: Minot 2021, Mondkar 2021, Prasantha 2019, and Kabir 2022 applied interventions across 3-4 building blocks but only Minot 2021 and Mondkar 2021 achieved increased coverage.
- Three of the nine studies (33%) that had interventions across one to two building blocks attained increased coverage. Of the rest nine studies, only three studies, i.e., Kapoor 2021, Joshi 2022 and Hendricks-Munoz 2014 achieved increased coverage.

The studies that did not achieve increased coverage had interventions primarily focused on the health workforce and service delivery and were weak on leadership and governance, financing, and health information systems.

Studies that included high-intensity intervention in at least one health system building block were more likely to achieve increased KMC coverage than those that did not:

- All three studies (100%) that achieved increased KMC coverage along with mean skin-to-skin contact for ≥8 hours per day had high-intensity interventions in at least one health system building block.
- Three of the five studies (60%) that achieved an increased KMC coverage but where mean skin-to-skin contact was < 8 hours per day or not reported had high-intensity interventions in at least one health system building block.
- Three of the eight studies (38%) studies that did not achieve an increased KMC coverage had high-intensity interventions in at least one health system building block.

The type of interventions implemented by the studies that achieved increased KMC coverage are summarized in Table 6.

**Table 6.** Summary of key interventions required to achieve increased KMC coverage.

| **HS BBs** | **Key interventions** |
| --- | --- |
| **Leadership and Governance or Policy** | 1. High-level leadership engagement<br>2. KMC supportive policies & regulations on staff licensing and facility standards |
| **Health workforce** | 3. Ensure adequate nursing staff with strengthened competency and motivation to support KMC<br>4. Engagement of professional organizations, KMC ‘champions’, maternal-neonatal staff collaboration |
| **Health financing** | 5. Govt. funding of KMC wards, human resources, running costs<br>6. Expanded health insurance for small babies |
| **Service delivery** | 7. KMC wards with a conducive environment (infrastructure, support, and counselling)<br>8. Community engagement to promote KMC<br>9. Early identification and facilitated referral of LBW babies |
| **Health information systems** | 10. Recording KMC (clinical registers) using KMC-specific indicators in routine data systems |
| **Supplies** | 11. KMC beds, chairs, and garments |

## DISCUSSION

The review findings suggest that a higher proportion of studies that applied interventions across more health system building blocks achieved increased KMC coverage-100% studies that applied interventions across 5-6 building blocks, 50% studies that applied interventions across 3-4 building blocks, and 33% studies that applied interventions across 1-2 building blocks achieved increased coverage. The studies that did not achieve increased coverage had interventions primarily focused on the health workforce and service delivery and were weak on leadership and governance, financing, and health information systems. Also, studies that included high-intensity intervention in at least one health system building block were more likely to achieve increased KMC coverage than those that did not. The type of interventions implemented in studies that achieved a large increase in coverage are summarized.

KMC is a complex intervention, both as individual practice requiring specific behaviours from the infant’s mother and family to carry and feed their preterm or LBW infant for a prolonged period and as a programmatic component of small and/or sick newborn care wherein the health systems need to be ready to provide essential support to enable the mother and family to practise KMC. We found only one study looking at demand-side intervention of financial incentives to mothers (37) which was not eligible for inclusion, none related to improving care seeking practices or access to care though one study did address it through community interventions to identify and refer LBW babies (25).

Few studies applied supply-side health interventions and reported KMC coverage. Those that did were quite variable in several aspects yet do allow some useful interpretations. System wide changes are not possible without coordinated and complementary actions in multiple domains. However, some health system actions in the domain of leadership and governance, health financing and health information systems may be particularly more effective alongside service delivery improvement and health workforce interventions, esp. to achieve increased KMC coverage at scale.

Effective leadership and management have been shown to lead to optimal performance of health systems and positively impact health services and the population’s health outcomes (38), and changes in policy environment and availability of a strategic plan for scale-up have been shown to facilitate scale up (39). However, the level of engagement, intensity and sustenance of intervention in this domain appears to be an important factor influencing coverage, esp. at the national and sub-national scale, as evidenced by the two studies that achieved high coverage and one study that achieved low coverage but where this was the primary high-intensity intervention, potentially a major factor responsible for the observed change in coverage (23-25).

Health financing is another crucial aspect. Only two of the eight studies that achieved high coverage reported health financing as an interventions, but it is noteworthy that both were large scale studies and achieved improvement in coverage at the district and national level (24,25). This suggests that while coverage improvements at facility-level may be achieved with local resources, programmatic scale up of KMC will require dedicated health system financing.

Next health system building block is the health information systems. All three studies that achieved high KMC coverage with high SSC duration had interventions in the health information systems (25,26,35). It is obvious that prolonged and continuous KMC cannot be achieved without adequate routine monitoring. Only two of the five other studies that achieved high coverage intervened in this domain, but it is interesting to note that the remaining three studies already had high KMC coverage at baseline (>75%), which means they may already have strong KMC measurement and monitoring in place.

Health work force interventions in isolation had little effect (32,33,34) as also identified in the past (40,41). Service delivery interventions varied by context, but key factors that facilitated prolonged and continuous SSC involved infrastructure changes to keep mothers and families with their infants too practice KMC, providing them the required support and counseling along with family involvement. Supplies for KMC, i.e., beds, chairs and garments are necessary but are all low-cost items.

The findings of the review imply that a systems wide approach guided by leadership and good governance with few key set of health system actions can help increase KMC coverage at scale and is feasible to do in most settings.

Our review is perhaps the first attempt to objectively evaluate the health system interventions that may improve KMC coverage. It spans national-level implementation through district and facility levels, with findings that are consistent with systematic reviews on barriers and enablers on KMC implementation (42,43), as well as learnings from programmatic implementation of KMC (44). However, there are several potential limitations to acknowledge. Publication bias is likely as studies which applied interventions to improve KMC but did not achieve expected or desired coverage may not have been published, and it could not be assessed in this review. The included studies were heterogenous in terms of setting, population, methods, scale, intensity and quality of intervention delivery, and time of outcome measurement, though the qualitative analysis did attempt to categorize the studies and interpret the findings using an objective framework. Owing to the nature of the review, results cannot be attributed to individual components of the intervention packages and must be considered together as package of health system building blocks.

To conclude, high-intensity interventions across multiple health system building blocks should be used for equitably scaling up KMC. High-intensity interventions across multiple health system building blocks should be used for equitable scale-up of KMC.

## Data Availability

All data produced in the present study are available upon reasonable request to the authors.

https://www.crd.york.ac.uk/PROSPEROFILES/271834_STRATEGY_20210804.pdf

## Acknowledgement

We acknowledge Dr. Rajiv Bahl, formerly, Head of the Newborn Health unit and Head of the Newborn Maternal, Newborn Child and Adolescent Health Research, Department of Maternal, Newborn, Child, Adolescent Health and Ageing, World Health Organization, Geneva, Switzerland; and currently, Secretary, Department of Health Research, Government of India, and Director General, Indian Council of Medical Research, India, for his advice on data synthesis and interpretation.

